# Diverse outcomes of controlled human malaria infection originate from host-intrinsic immune variation and not *var* gene switching

**DOI:** 10.1101/2020.09.04.20188144

**Authors:** Kathryn Milne, Alasdair Ivens, Adam J. Reid, Magda E. Lotkowska, Áine O’Toole, Geetha Sankaranarayanan, Diana Muñoz Sandoval, Wiebke Nahrendorf, Clement Regnault, Nick J. Edwards, Sarah E. Silk, Ruth O. Payne, Angela M. Minassian, Navin Venkatraman, Mandy Sanders, Adrian V.S. Hill, Michael P. Barrett, Matthew Berriman, Simon J. Draper, J. Alexandra Rowe, Philip J. Spence

## Abstract

Falciparum malaria is clinically heterogeneous and the relative contribution of parasite and host in shaping disease severity remains unclear. We explored the interaction between inflammation and parasite variant surface antigen (VSA) expression, asking whether this relationship underpins the variation observed in controlled human malaria infection (CHMI). We uncovered marked heterogeneity in the response of naive hosts to blood challenge; some volunteers remained quiescent, others triggered interferon-stimulated inflammation and some showed transcriptional evidence of myeloid cell suppression. Significantly, only inflammatory volunteers experienced hallmark symptoms of malaria. When we tracked temporal changes in parasite VSA expression to ask whether variants associated with severe disease preferentially expand in naive hosts (as predicted by current theory) we found that *var* gene profiles were unchanged after 10-days of infection. The diverse outcomes of CHMI therefore depend upon human immune variation and there is no evidence for switching or selection of *var* genes in naive hosts.

## Introduction

There is enormous diversity in the human response to identical immune challenge. This variation is currently being interrogated in large cohorts of healthy volunteers to pinpoint the genetic and non-genetic factors that dictate human immune decision-making (1–6). Controlled human malaria infection (CHMI) provides a well-established challenge model in which to examine immune variation *in vivo*, and heterogeneity in the host response to *Plasmodium falciparum* is well-recognised (7,8). Prior work suggests a dichotomy in immune decision-making with some individuals triggering an inflammatory response, which is characterised by high circulating levels of IFNγ, whilst others mount an alternative response distinguished by the early release of TGFβ and suppression of IFNγ signaling (9–12). The precise mechanisms underpinning these divergent responses remain unknown.

Genome-wide transcriptional profiling has the potential to map immune decision-making in unprecedented detail and previous CHMI studies have revealed an upregulation of genes associated with pathogen detection, NFκB activation and IFNγ signaling consistent with a systemic inflammatory response (13–15). A transcriptional signature of an alternative immune response has not been described in naive hosts. Nevertheless, these prior studies were primarily designed to measure shared or common signatures of infection, potentially masking variation between volunteers. Only one study (16) has specifically investigated transcriptional variation in CHMI and it reported two distinct outcomes based on host microRNA profiles; notably, the authors were able to correlate the induction of 3 specific miRNAs with a lower parasite burden suggesting that events early in infection may have important downstream consequences. Identifying the sources of immune variation may therefore reveal some of the factors that underpin the clinical heterogeneity of falciparum malaria. In this context, direct intravenous challenge with blood-stage parasites provides a unique opportunity to assess the contribution of host-intrinsic mechanisms of variation as it ensures all volunteers receive an identical immune challenge (17).

The potential role of parasite factors in influencing human immune decision-making has not yet been explored in CHMI. Expression of a subset of parasite variant surface antigens (VSA) known as group A and DC8 *var* genes is associated with severe malaria in both children and adults (18–23). These virulence-associated genes encode PfEMP1 adhesion molecules that mediate sequestration of infected red cells in the microvasculature, contributing to pathology. Group A/DC8-expressing parasites are thought to be more pathogenic than those expressing other *var* types (group B and C) and can bind the endothelium with high affinity in sensitive sites, such as brain (24). It is unknown whether expression of these variants can influence the immune response, for example by causing dysregulated activation that leads to widespread collateral tissue damage. Or conversely, whether inflammation might preferentially support the survival and expansion of these variants by upregulating their binding sites on the endothelium.

It has been observed in previous CHMI studies that parasites transcribe a broad array of *var* genes in naive hosts, with predominant expression of group B variants and no marked differences between volunteers (25, 26). Indeed, we have shown that mosquitoes reset malaria parasites to ensure diverse expression of their VSA repertoire at the start of the blood cycle (27). It therefore remains unclear how group A and DC8 *var* genes seemingly come to dominate the PfEMP1 landscape in severe malaria. Antibody-mediated clearance of parasites expressing group B variants is a possible explanation, but is unlikely because severe malaria develops in hosts with low or absent immunity during their first few infections of life (28). It has instead been suggested that parasites expressing group A/DC8 *var* genes preferentially expand in naive hosts because they sequester and avoid host clearance more effectively than parasites expressing group B or C *var* genes (29–31). This explanation is widely accepted but is not based on substantial evidence.

In this study, we have used a blood challenge model to investigate host-intrinsic variation in the immune response to falciparum malaria and examined the interplay between parasite and host factors in shaping outcome of infection. This model has the advantage that an identical immune challenge is given to all volunteers and furthermore the frequency of each parasite variant can easily be measured at the start and end of infection to track changes through time. We set out to test the specific hypotheses that **a**, expression of group A and DC8 *var* genes would preferentially increase as the infection progressed and **b**, there would be a measurable relationship between inflammation and parasite VSA expression, which would shape the clinical outcome of infection.

## Results

### Immune decision-making in falciparum malaria

To explore host-intrinsic variation in the immune response to *P. falciparum* a cohort of 14 malaria-naive volunteers were infected with an equal number of blood-stage parasites (clone 3D7) by direct intravenous inoculation (Table S1). This enabled the pathogenic blood cycle to be initiated in every volunteer within a 30-minute window (32). Systemic changes in the host response were then captured through time by transcriptionally profiling whole blood every 48-hours from the day before infection until the day of diagnosis (the point of drug treatment). This time-point varied between volunteers, occurring 7.5 to 10.5 days post-infection when two out of three diagnostic criteria were met (positive thick blood smear and/or parasite density > 500 parasites ml^−1^ by qPCR and/or symptoms consistent with malaria). To reveal the diversity of responses within our cohort each volunteer’s time-course was analysed independently by tracking their dynamic changes in gene expression as the infection progressed. Importantly, this approach does not assume shared features between individuals, nor does it bias against uncommon (or rare) responses.

As a first step, we measured the variance of every gene through time and visualised the 100 protein-coding genes with highest variance in each volunteer (Fig. S1). To control for baseline variation in gene expression, uninfected control volunteers were also analysed. We found that variance in one third of infected volunteers (4/14) was comparable to the uninfected control group, suggesting these four volunteers did not respond to infection (Table S2). To explore this further, we merged the top 100 gene lists from all infected volunteers to produce a single non-redundant list of the most dynamically expressed protein-coding genes across the cohort (n = 517 unique genes, henceforth called the 517-gene superset); we could then directly compare the transcriptional response between individuals. Accordingly, we performed a principal component analysis (PCA) of these genes through time, and for each volunteer we determined the Euclidean distance travelled to quantify the magnitude of their response (Fig. S2 and Table S2). Using the variance and distance travelled metrics we then set two thresholds to identify volunteers that trigger a measurable response to blood challenge (see methods); four volunteers failed to cross both thresholds (v018, v012, v208 & v020) and were therefore grouped and labelled unresponsive. A pairwise comparison between their diagnosis and pre-infection samples confirmed that there were zero differentially expressed genes in this group (adj p < 0.05). Immune quiescence is therefore a common outcome of controlled human malaria infection.

In contrast, variance (Fig. S1) and distance travelled (Fig. S2) both suggested that the remaining 10 volunteers made a robust response to infection (Table S2). And when we clustered and visualised the 517-gene superset expression data we found that these volunteers could separate easily into two groups according to whether they up- or downregulated gene expression at diagnosis (Fig. 1 and Table S3). Based on this observation, we grouped the 8 volunteers who upregulated gene clusters 2 & 4 and performed a pairwise comparison between their diagnosis and pre-infection samples; we uncovered 2,028 differentially expressed genes (adj p < 0.05) (Table S4). To gain insight into the biological functions of this response we used ClueGO (33, 34) to first identify the significant GO terms associated with these genes and then place them into a functionally organised non-redundant gene ontology network. ClueGO uncovered 217 GO terms across 34 functional groups (Table S5); the most significant groups related to interferon signaling, activation of myeloid cells, production of inflammatory molecules and activation of T cells (Fig. 2a-b). These terms all indicate systemic interferon-stimulated inflammation (a well-described host response to malaria in mouse and human studies (35–37)) and we therefore labelled this group inflammatory. We then biologically validated ClueGO’s in silico finding by measuring the plasma concentrations of two interferon-stimulated chemokines, CXCL9 and CXCL10. These chemokines are primarily produced by myeloid cells and specifically function to recruit and activate T cells (38). We found a > 2-fold increase in one or both of these chemokines at diagnosis in every volunteer in this group but no other volunteer (Fig. 2c).

**Fig. 1.**
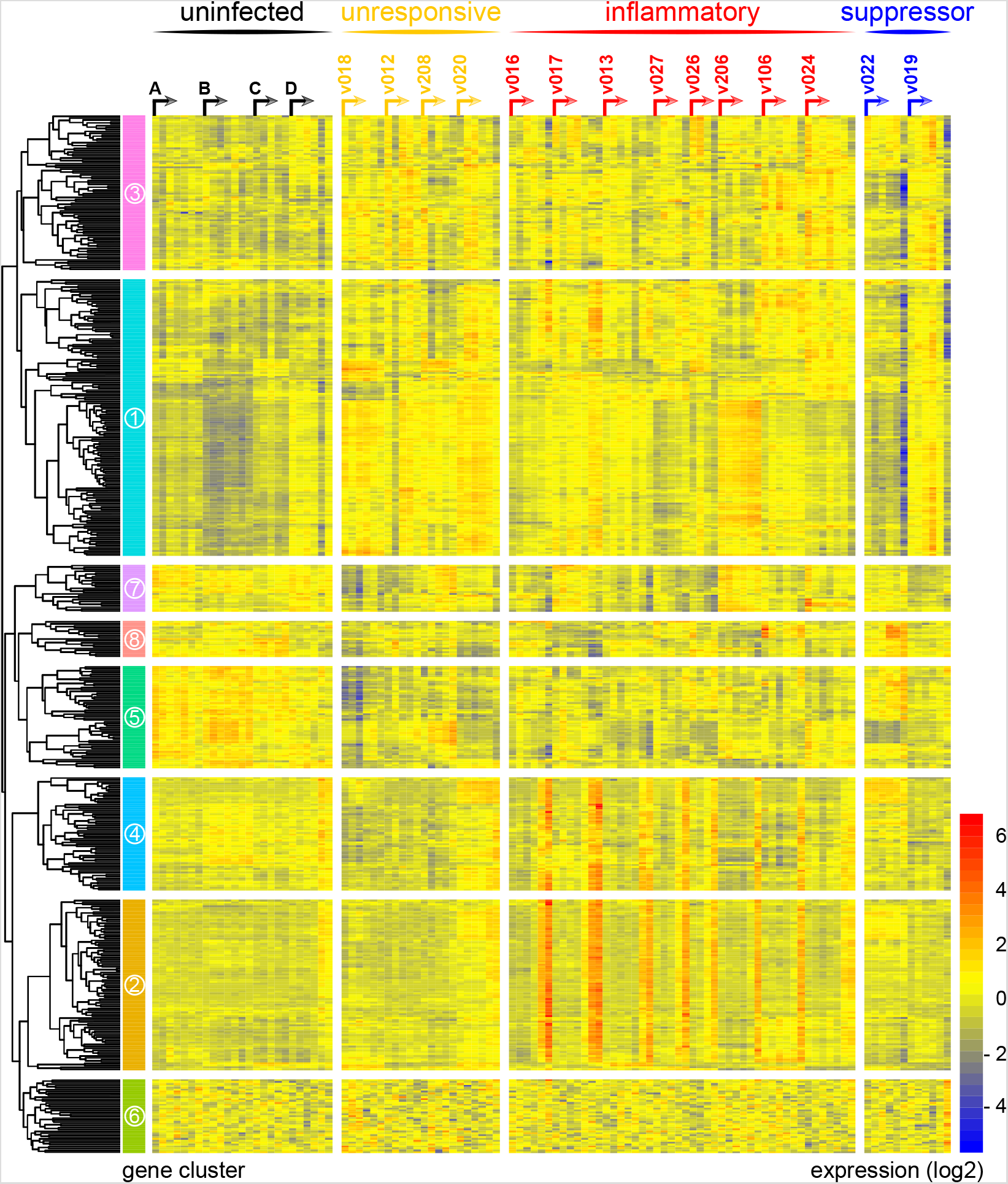
An identical immune challenge leads to diverse outcomes in falciparum malaria. Log2 expression values of 517 protein-coding genes in whole blood during infection. Genes (rows) are ordered by hierarchical clustering, whereas whole blood samples (columns) are ordered by volunteer and time-point (pre-infection to diagnosis, left to right). Arrows start from the pre-infection sample and volunteers are grouped by host response (refer to methods). Uninfected controls demonstrate minimal within-host variation in expression of these genes. Median sample number per volunteer = 6.

**Fig. 2.**
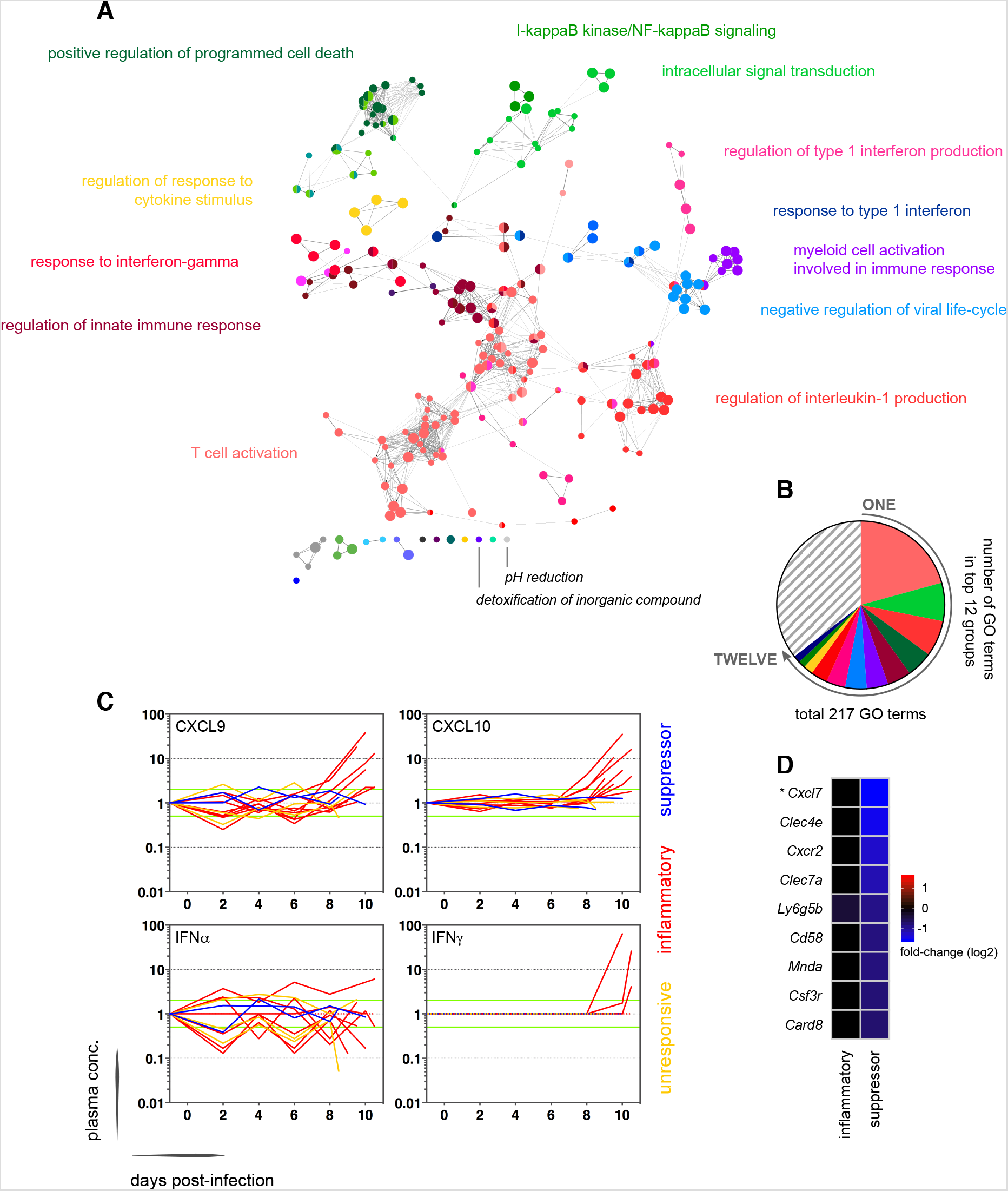
Interferon-stimulated inflammation is the leading outcome of immune decision-making. (**A**) Gene ontology network of the 2,028 genes differentially expressed at diagnosis in the inflammatory group. Each node represents a significantly enriched GO term (adj p < 0.05) and node size is determined by significance (bigger nodes have lower p values). Nodes are interconnected according to their relatedness (kappa score >0.4) and grouped if they are connected and share > 40% genes. Each functional group is then given a unique colour and the leading GO term in the top 12 groups is highlighted. Two GO terms of interest, which are not part of any functional group, are also shown in italics. (**B**) The proportion of GO terms in each of the top 12 functional groups; collectively, these account for two thirds of all significantly enriched GO terms in inflammatory volunteers. (**C**) Plasma concentration of interferon alpha & gamma and interferon-stimulated chemokines (CXCL9 & CXCL10) during infection. One line represents one volunteer (no data for v020) and lines are colour-coded by host response. For each volunteer, all data points are normalised to their own baseline (day –1); horizontal green lines represent a 2-fold increase or decrease compared to baseline. (**D**) Log2 fold-change of 9 immune genes involved in myeloid cell differentiation, activation and function in whole blood at diagnosis. Data are presented relative to pre-infection samples and all genes are significantly downregulated in the two suppressor volunteers (adj p < 0.05). \**Cxcl7* is the common gene name for pro-platelet basic protein *(Ppbp)*.

The final two volunteers were also grouped (and given the suppressor tag) based on their apparent downregulation of clusters 1 & 3 at diagnosis (Fig. 1). Significantly, a pairwise comparison between their diagnosis and pre-infection samples revealed a core signature of 77 genes that were differentially expressed in response to blood challenge (adj p < 0.05) (Table S6). The majority of these genes (62/77) were downregulated and crucially > 85% were unique to this group, including key regulators of monocyte & neutrophil differentiation *(Csf3r)* and recruitment & activation *(Cxcl7, Cxcr2, Clec4e, Clec7a* and *Mnda)* (Fig. 2d). A pairwise comparison between the inflammatory and suppressor groups confirmed that all of these six immune genes were differentially expressed at diagnosis, together with a further 887 genes (adj p value < 0.05) (Table S7). Evidently, these host response profiles do not overlap and instead likely represent distinct *in vivo* outcomes of human immune decision-making. Nevertheless, a much larger sample size would be required to fully elucidate immune suppression as an alternative response to *P. falciparum*.

Collectively, these data reveal at least three possible early outcomes of immune decision-making in falciparum malaria – immune quiescence, interferon-stimulated inflammation and suppression of myeloid-associated gene expression.

### Sporozoites have a limited role in shaping the human response to blood-stage infection

We next explored the influence of the pre-erythrocytic-stages of infection on the human immune response to blood-stage parasites. Whilst blood challenge uses a recently mosquito-transmitted parasite line (< 3 blood cycles from liver egress (39)) this model nevertheless bypasses the skin and liver stages, which have been proposed to elicit regulatory mechanisms that could modify the subsequent immune response to blood-stage infection (40). We therefore inoculated 5 additional malaria-naive volunteers with the same parasite genotype but this time by the natural route of infection, mosquito bite (Table S1). As before, we transcriptionally profiled whole blood from the day before infection until diagnosis (day 11), including the final day of liver-stage infection (day 6). And once more, every volunteer was analysed independently this time using EdgeR to identify highly dispersed genes within each time-course (see methods). After merging these lists, we produced a single non-redundant list of the most dynamically expressed genes across the mosquito challenge cohort (n = 117 unique genes, henceforth called the 117-gene superset) (Table S8). When we clustered and visualised the expression data within this superset we found that 116 out of 117 genes were upregulated exclusively during the blood cycle (Fig. S3a-b). And when we examined the biological functions of these genes ClueGO identified response to interferon gamma (GO: 0034341) as the most significantly enriched GO term (Table S9). Notably, this was also the most significant GO term in inflammatory volunteers infected by blood challenge (Table S5), indicating concordance between the two infection models. In agreement, 19 of the 21 genes that display the highest fold change in inflammatory volunteers (blood challenge) are also upregulated in the 117-gene superset (mosquito challenge) (Table S8). And at protein level, high circulating levels of CXCL10 are detectable at diagnosis in four of the five volunteers infected by mosquito bite (Fig. S3c). Our data therefore demonstrate that interferon-stimulated inflammation is the dominant human response to blood-stage parasites regardless of the route of infection. Furthermore, our data fail to provide compelling evidence of a systemic transcriptional response to liver-stage infection in human malaria (Fig. S3b). Taken together, these findings indicate that the pre-erythrocytic-stages of infection may have a limited role in shaping the immune response during the pathogenic blood cycle.

### Systemic inflammation coincides with the onset of clinical symptoms

We therefore moved on to investigate the factors that influence immune decision-making in the blood challenge model and the consequences of these divergent paths. In the first instance, we asked whether the rate of parasite replication could shape the host response to infection; after all, a simple assumption might be that inflammation is triggered by fast-growing parasites. The parasite multiplication rate (PMR, modelled from the qPCR data (32)) varied considerably between volunteers (range 6.7 - 12.8) despite every individual receiving an equal number of infected red cells (Table S1). However, the parasite multiplication rate of the inflammatory group was not significantly different to that of unresponsive or suppressor hosts (Fig. 3a-b) and therefore appears to have little bearing on the outcome of human immune decision-making. Inflammatory volunteers did, however, tend to have a longer course of infection (Fig. 3a) and hence the parasite burden at diagnosis was higher for the inflammatory group (median 30,309 parasites ml^−1^, range 9,133 to 273,247) than the rest of the cohort (median 7,899 parasites ml^−1^, range 1,440 to 19,670) (Mann Whitney test, p = 0.02) (Table S1). Parasite burden may therefore promote an inflammatory response in this challenge model but cannot fully explain it; for example, 2/4 unresponsive volunteers (v018 & v020) had parasite densities that were comparable to 4/8 inflammatory volunteers at diagnosis (v017, v027, v026 & v106) (Table S1). Our data therefore support previous observations that the parasite density threshold required to trigger inflammation is highly variable between individuals (41, 42).

**Fig. 3.**
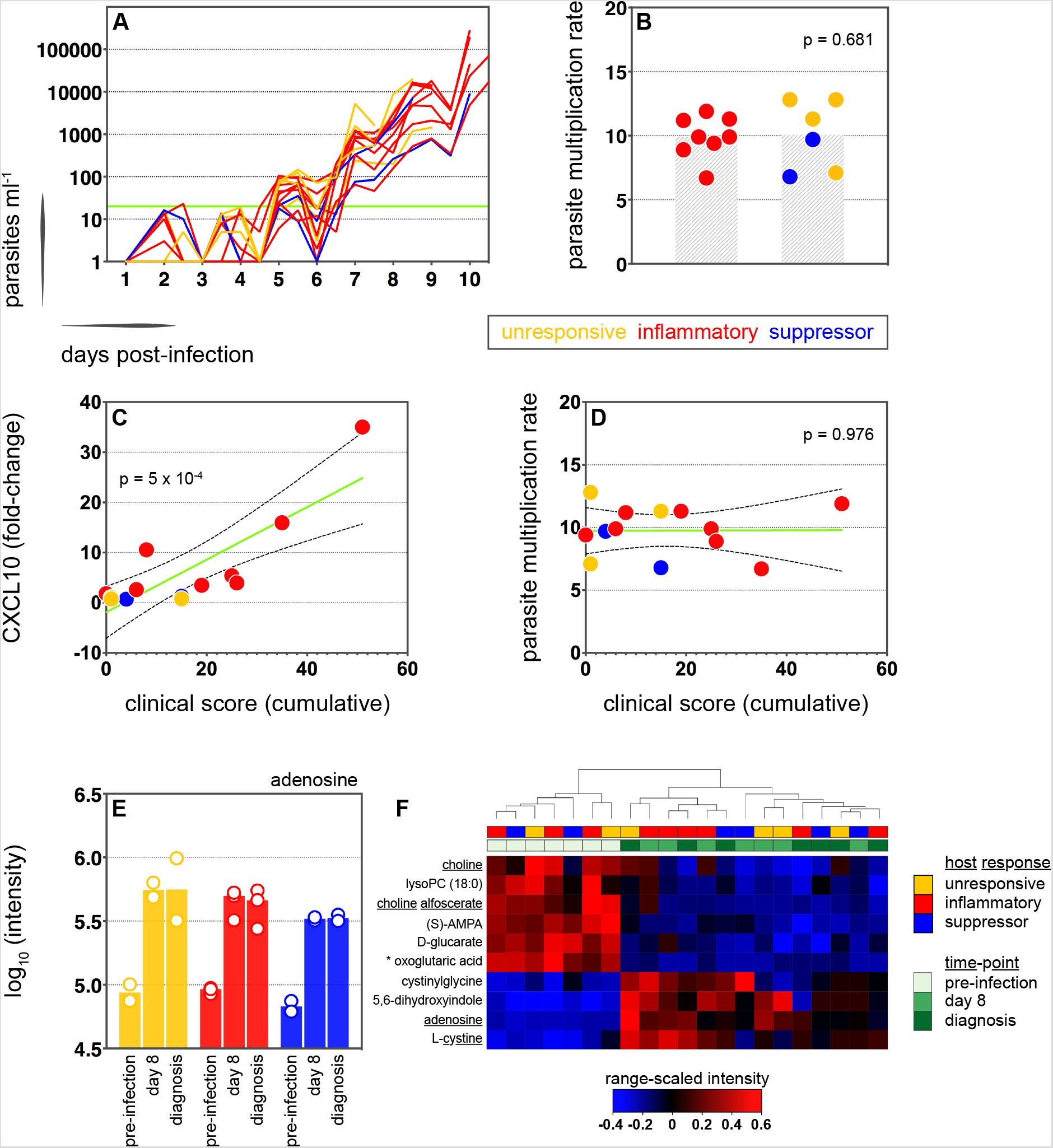
Symptoms scale with the intensity of inflammation. (**A**) Parasite growth curves colour-coded by host response; each line represents one volunteer. Blood samples were collected every 12-hours for qPCR analysis of circulating parasite density and the horizontal green line represents the lower limit of quantification (20 parasites ml^−1^). (**B**) Parasite multiplication rates colour-coded by host response; each dot represents one volunteer and shaded areas show the mean value. A Mann Whitney test was used to ask whether the parasite multiplication rate observed in the inflammatory group was different to all other volunteers (p value is shown). (**C** and **D**) Linear regression of CXCL10 (C) or parasite multiplication rate (D) plotted against clinical score (the sum of adverse events during infection). CXCL10 fold-change measures plasma concentration at diagnosis over baseline (day –1). One dot represents one volunteer (no data for v020) and dots are colour-coded by host response. The green line represents the best-fit model (p value of the slope is shown) and dashed lines are the 95% confidence intervals. (**E**) Log10 transformed intensity values of adenosine in plasma during infection. An authentic standard was run in tandem with all samples to validate adenosine detection. (**F**) Range-scaled intensity values of 10 plasma metabolites that were differentially abundant during infection. Metabolites (rows) and samples (columns) are ordered by hierarchical clustering. Note that an authentic standard was used to validate detection of all underlined metabolites and the full name for oxoglutaric acid is 4-hydroxy-2-oxoglutaric acid. (E and F) Only plasma samples from the most inflammatory and symptomatic volunteers (v016, v017 & v013), the suppressor volunteers (v022 & v019) and two unresponsive volunteers (v018 & v208) were analysed for metabolite abundance.

We next set out to ask whether the diverse outcomes of immune decision-making had clinical consequences. Symptomatology was scored throughout infection with adverse events graded as absent (0), mild (1), moderate (2) or severe (3) (see methods). Volunteers in the unresponsive and suppressor groups were essentially asymptomatic at diagnosis, whereas inflammatory volunteers exhibited more frequent and/or severe adverse events, including fever (Fig. S4 and Fig. S5a). When we then calculated a cumulative clinical score for each volunteer (by summing all adverse events throughout infection) we found a strong positive correlation between symptoms and the intensity of inflammation (Fig. 3c). In contrast, there was no correlation between clinical score and parasite multiplication rate (Fig. 3d). We then measured circulating levels of Angiopoietin-2, a biomarker of endothelium dysfunction and indicator of disease severity (43); only one volunteer displayed elevated levels of Angiopoietin-2 (Fig. S5b) and this individual was the most inflammatory host in the cohort (based on upregulation of plasma CXCL10 & IFNγ). What’s more, haematological analysis revealed that only inflammatory volunteers experienced acute lymphopenia, a conserved hallmark of clinical malaria (Fig. S5c-d) (44). Symptomatology is therefore not driven by the rate of parasite replication in controlled human malaria infection. Instead, clinical outcome seems to depend upon the intensity of inflammation, which varies widely between hosts.

In light of these findings, we examined the potential impact of inflammation on host metabolism to better resolve the consequences of immune decision-making. Here we used an LC-MS-based metabolomics platform to measure changes in circulating metabolites during infection, and applied range-scaling to ensure all metabolites (and their temporal fluctuations) were equally weighted (45). When we compared the three most inflammatory and symptomatic volunteers to unresponsive and suppressor hosts we could find no metabolic signature of inflammation that was robust to multiple testing (FDR-corrected p value < 0.1). On the other hand, if we simply considered all volunteers as a single group and compared their post-infection time-points (day 8 and diagnosis) against pre-infection samples we could identify a conserved and persistent signature of blood-stage infection (Fig. S6). This was characterised by increased adenosine (promotes vasodilation (46)), choline depletion (supports parasite membrane synthesis (47)) and reduced lysoPC (induces gametocytogenesis(48)) (Fig. 3e-f). Evidently, volunteers who are immune quiescent are nevertheless metabolically responsive to *P. falciparum*, and their response is indistinguishable from inflammatory hosts.

All together, these data reveal that immune decision-making, systemic metabolism and parasite replication all operate independently in the early stages of the blood cycle. And they further reveal a clear relationship between the intensity of inflammation and symptomatology in falciparum malaria.

### Parasite variants associated with severe disease do not expand in naive hosts

To investigate the origin of immune variation and gain further insight into the factors that can shape outcome of infection we measured parasite VSA expression. To this end, we applied ultra-low input RNA-sequencing methodology to parasites isolated during CHMI and mapped their transcriptional profiles from the start to end of infection. In the first instance, we examined the hierarchy of *var* gene expression in the inoculum used for blood challenge; this derived from a single malaria-infected volunteer infected by mosquito bite and was cryopreserved *ex vivo* (without culture) after just 3 cycles of asexual replication (39). Although each individual blood-stage parasite will only transcribe a single *var* gene (49) the parasite population as a whole can express many different variants within a host. In agreement, we found more than 20 *var* genes highly transcribed in the inoculum; notably, the expression hierarchy was dominated by group B variants and there was very low (or undetectable) transcription of virulence-associated group A or DC8 *var* genes (Fig. 4a and Table S10). The one exception was PF3D7_0400400, which encodes a group A brain endothelial cell binding PfEMP1 variant (50). We found that group B *var* genes also dominated the PfEMP1 landscape when we isolated parasites from our mosquito challenge cohort on days 9–11 post-infection (2-3 cycles after liver egress) (Fig. 4a and Fig. S7a-b). In these samples the proportion of *var* gene reads mapping to group A was higher than in the inoculum and the two DC8 variants were highly transcribed in some volunteers (Table S10). Nevertheless, these data confirmed previous observations that whilst malaria parasites exit the human liver expressing a diverse *var* gene repertoire variants associated with severe disease constitute a minority population (25, 26, 51, 52).

**Fig. 4.**
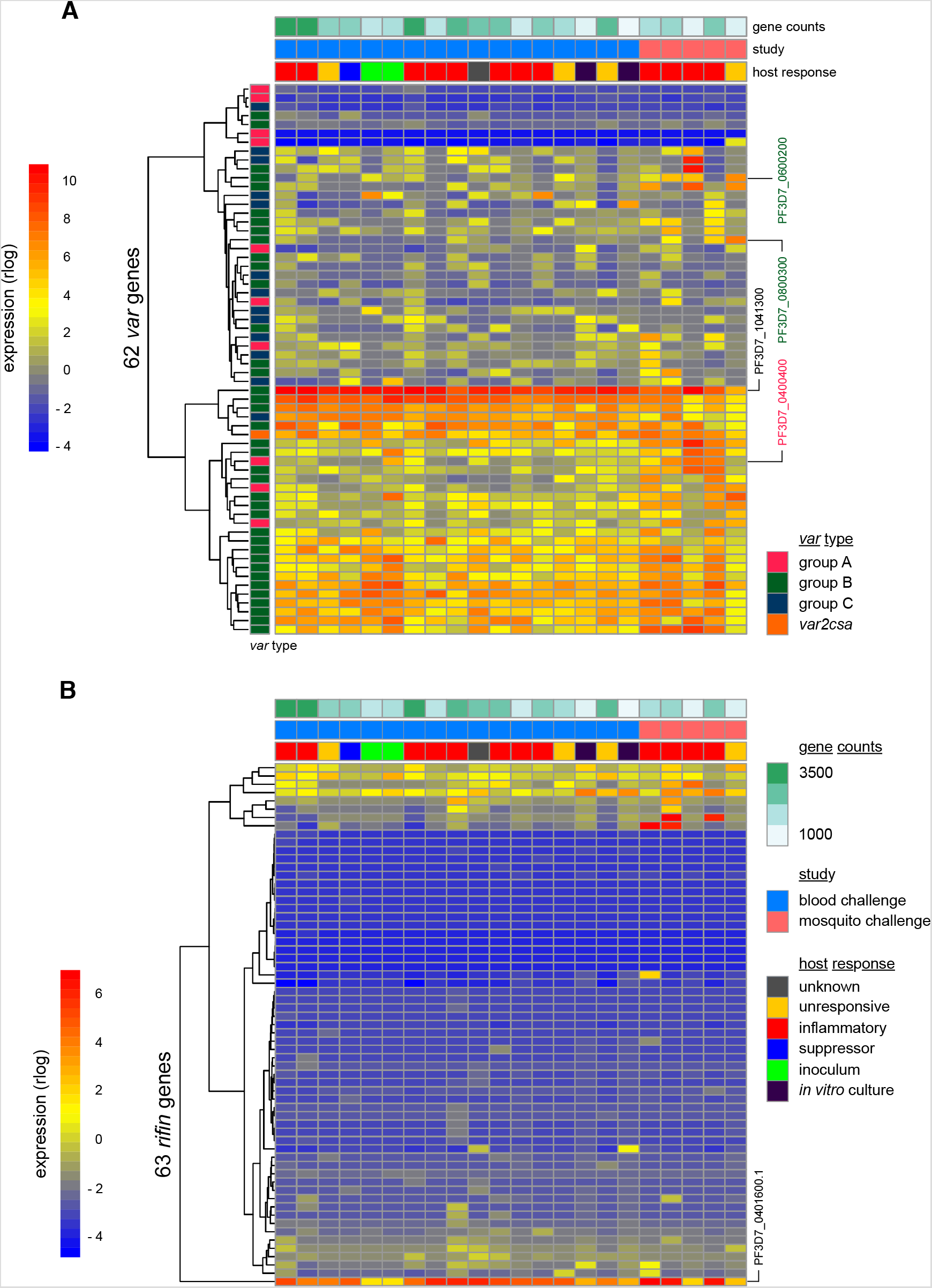
Switching & selection of variant surface antigens does not occur in naive hosts. (**A** and **B**) Rlog expression values of *var* (A) and *rifin* (B) genes in the inoculum and diagnosis parasite samples after blood challenge (blue study); samples obtained from the second & third blood cycle after mosquito challenge are also shown (pink study). Genes (rows) are ordered by hierarchical clustering and colour-coded by *var* type; parasite samples are colour-coded by host response; and *in vitro* cultured ring-stage parasites are shown for comparison. Two volunteers did not have parasite sequencing data (unresponsive volunteer v208 & suppressor volunteer v022) and the two inoculum samples are technical replicates of one biological sample. The *var* (PF3D7_1041300) and *rifin* (PF3D7_0401600) genes dominantly expressed across all samples are labelled. *Var* genes associated with severe disease (group A variant PF3D7_0400400 and DC8 variants PF3D7_0800300 & PF3D7_0600200) are also labelled. Gene counts show the number of parasite genes that have at least 3 uniquely mapping reads; this provides a measure of genome coverage in every sample.

Current theory predicts that group A and DC8 variants should preferentially expand in naive hosts, either by transcriptional switching or by more effective sequestration (and improved survival) favouring their selection (29–31). Blood challenge provides a unique opportunity to directly test this hypothesis as it increases the number of asexual replication cycles over which variant switching & selection can take place (cf. mosquito challenge (53)). Parasites were therefore isolated from volunteers infected by blood challenge and processed immediately (without culture) for RNA-sequencing; after 6 cycles of replication *in vivo*, the expression hierarchy of *var* genes at diagnosis was essentially unchanged from that observed in the inoculum (Fig. 4a and Fig. S7b). For example, the group B *var* gene PF3D7_1041300 remained the most highly expressed variant in every volunteer, and transcription of group A and DC8 *var* genes remained low or undetectable (Table S10). Surprisingly, expression of the brain endothelial cell binding variant PF3D7_0400400, which was transcribed in the inoculum, was decreased at the end of infection in every volunteer (Fig. 4a). We could further highlight the significance of this result by looking only at intron-spanning reads, which must derive from functional *var* gene transcripts (rather than regulatory antisense transcripts) (54). We found that PF3D7_0400400 was functionally transcribed in the inoculum but was entirely absent in all volunteer samples (Fig. S7c). Differential gene expression analysis (using DESeq2) then confirmed that no *var* gene was more highly transcribed in volunteers than in the inoculum (adj p < 0.01). Evidently, there was no preferential expansion of group A or DC8 variants *in vivo* in this study.

When we expanded our analysis to include *rifin* genes we still could find no evidence of gene switching or selection over the course of infection. Indeed, only a single *rifin* (PF3D7_0401600) was differentially expressed in the volunteer samples compared to the inoculum (log2 fold-change = 7.59, adj p = 0.0002) and this gene was already the dominant *rifin* at the start of infection (Fig. 4b). It should be noted, however, that although *ex vivo* RNA-sequencing is ideal for *var* gene analysis (as blood samples contain mostly ring-stage parasites) it is less suited to *rifins*, which are predominantly transcribed at later stages of the parasite life cycle (55). We therefore moved on to directly test the relationship between inflammation and parasite VSA expression. Once again, we used DESeq2 to identify differentially expressed parasite genes but this time between inflammatory volunteers and the rest of the cohort. There were no differentially expressed *var* or *rifin* genes (adj p < 0.01). In fact, only one differentially expressed gene could be identified (PF3D7_1136500, *Pfck1)* - this gene encodes casein kinase 1 and was more highly transcribed in the inflammatory group (log2 fold-change = 7.32, adj p = 0.002). The diverse clinical outcomes of CHMI cannot therefore be easily explained by variation in parasite gene expression; instead, they must largely originate from host-intrinsic immune variation.

## Discussion

Our data emphasise the importance of the human host in determining their own fate. And this is well supported by epidemiological studies showing that the clinical manifestations of severe malaria depend upon host age (56) and children learn to tolerate high parasite densities, which could otherwise cause life-threatening disease (28, 57). As such, we must understand the rules of immune decision-making if we want to uncouple mechanisms of pathology versus tolerance. In this study we set out to explore variation in the immune response to falciparum malaria and identified at least three possible early outcomes of blood-stage infection. The most likely outcome in a naive host is interferon-stimulated inflammation, which is characterised by activation of myeloid cells and systemic release of inflammatory molecules. Notably, only inflammatory volunteers develop hallmark symptoms of malaria (e.g. pyrexia and lymphopenia) providing further support for the idea that inflammation causes the earliest manifestations of disease. It is often assumed that the flip-side to this cost is immediate or early control of parasite burden and yet we find that the parasite multiplication rate of the inflammatory group is comparable to all other volunteers. It therefore appears that we might need to re-assess the role of innate inflammation in the first hours and days of the pathogenic blood cycle – rather than serving as an early brake on parasite replication it may be that the main purpose of this response is to switch bone marrow production in favour of myelopoiesis and mobilise the required effector cells to the spleen.

Recent evidence suggests that blood-stage parasites first trigger interferon signaling in the bone marrow (36) and the transcriptional changes we observe in whole blood are thus likely to be a signature of activated monocytes and neutrophils trafficking from bone marrow to spleen. Furthermore, the major secreted products of this emergency myeloid response (CXCL9 and CXCL10) specifically recruit T cells out of the circulation (38). The early innate response to malaria may therefore function primarily to re-structure and co-localise key strands of the immune system. Whether this leads to an effective response that can limit parasite burden and minimise collateral damage would then depend upon the outcome of cell-cell circuits in the remodelled spleen. Measuring the activation and differentiation of CD4^+^ T cells – the key orchestrators of innate and adaptive immunity – would provide a downstream readout of these critical tissue-specific interactions in human volunteers. Unfortunately, we can’t begin to examine these outcomes in our current datasets because at our final time-point (diagnosis) CD4^+^ T cells are still trapped in the spleen. Nevertheless, future studies could extend sampling beyond drug treatment to analyse resolution of the acute phase response and examine the heterogeneity between volunteers in T cell profiles as they are released back into the circulation. Due to the inherent adaptive plasticity of human T cells (58) this is likely to reveal much greater variation between hosts in their response to a first malaria episode.

A second common outcome in CHMI is immune quiescence. Although the course of infection tended to be shorter in unresponsive volunteers than the others this could not explain their lack of response to blood challenge. After all, half of unresponsive volunteers (2/4) had parasite densities comparable to half of the inflammatory group (4/8). And furthermore, v017 triggered a measurable inflammatory response at just 157 parasites ml^−1^ whereas v018 remained unresponsive at 20,000 parasites ml^−1^. These observations are entirely in-line with meta-analysis of historical malariatherapy data that measured the pyrogenic threshold in falciparum malaria as spanning more than 4-orders of magnitude (42). Human immune variation therefore dictates how you respond and when. Pushing up the pyrogenic threshold to disarm emergency myelopoiesis is a well-recognised host adaptation that promotes immune quiescence and allows individuals to transition from clinical to asymptomatic malaria (usually in adolescence) (41, 59). This is helpful if you have developed partial humoral immunity that can restrict total pathogen load but whether a pre-existing high threshold is advantageous in a naive host is unclear. This could indicate a natural tendency towards disease tolerance (60) or it may simply delay the time until an emergency myeloid response inevitably has to be kicked off to control parasite replication. Indeed, it remains a possibility that all naive hosts (including those that initially suppress genes involved in myeloid cell activation and recruitment) will eventually converge on interferon-stimulated inflammation as the infection progresses. After all, every volunteer increased their circulating levels of adenosine as part of a shared metabolic response to blood-stage infection and adenosine is a potent paracrine inhibitor of inflammation (61). This metabolic switch may therefore represent a hardwired mechanism of self-regulation because systemic inflammation is an unavoidable fate in naive hosts. Human challenge models could reveal the downstream consequences of early immune decision-making if they extend the window over which host fate can be tracked, and this can safely be achieved by increasing the parasite density at which asymptomatic volunteers need to be drug treated.

Our study also examined the *in vivo* relationship between inflammation and parasite VSA expression, asking whether variant switching and selection could shape clinical outcome during CHMI. We confirmed the previous finding that group B *var* genes, which encode PfEMP1 variants that are associated with uncomplicated malaria (62), are mainly expressed in human volunteers infected by mosquito bite (25, 26, 51, 52). The unique features of the blood challenge model then allowed us to examine the widely-held belief that group A and DC8 variants preferentially expand in naive hosts. Unexpectedly, we found only minor changes in *var* gene transcription over 6 cycles of asexual replication, with no evidence to support the expansion of virulence-associated variants *in vivo*. How then do group A and DC8 become the predominantly expressed *var* genes in parasites collected from severe malaria patients in endemic regions (18–23) and non-immune travellers (63, 64)? It is possible that switching or selection towards group A/DC8 variants occurs later in infection. However, this would have to occur rapidly because at the mean parasite multiplication rate of 10 seen in this study only 2–3 more cycles of replication would be needed for some volunteers to reach parasite burdens at which severe disease can occur (∼ 5,000 parasites ml^−1^ or 0.1% parasitaemia (65)). *Var* gene switching rates of culture-adapted parasites *in vitro* are low (estimated mean on-rates of 0.025–3% per generation (66–68)) and although mathematical modelling suggests the possibility of higher switching rates *in vivo* (69) experimental data are lacking.

Another possibility is that host inflammation leads to selection of parasites expressing group A and DC8 variants because cytokine-mediated upregulation of endothelial receptors (such as ICAM-1) increases sequestration efficiency and reduces their clearance, as compared to parasites expressing group B and C variants that bind only CD36 (70). However, in our study there was no evidence for increased group A/DC8 *var* gene expression in volunteers who showed clear clinical, transcriptional and biochemical evidence of inflammation. This study and most previous CHMI trials have used the 3D7 *P. falciparum* clone, or its parent line NF54 (71). Is it plausible that other parasite genotypes might predominantly express group A/DC8 *var* genes in naive hosts and therefore be more representative of parasites causing severe malaria in an endemic setting? The 3D7 parasite was cultured for many years prior to preparation of the inoculum (39) and could have acquired mutations (72) affecting its behaviour *in vivo*, or even reducing its ability to switch. However, in our mosquito challenge cohort there was considerable variation between volunteers in which group A and DC8 *var* genes were highly transcribed at the start of the blood cycle indicating that 3D7 has retained its potential to express a wide range of virulence-associated variants. And moreover, in a previous study parasites collected from volunteers given the same 3D7 inoculum as that used here were shown to switch when returned to culture (51). We also note a recent CHMI study using the 7G8 *P. falciparum* clone showed mostly group B *var* genes expressed in human volunteers (25). Existing data do not therefore support the idea of parasite genotype-specific differences in *var* gene expression in naive hosts, but more studies with diverse parasite genotypes are needed.

To complement the study of *var* gene switching and selection in naive hosts future work should also aim to examine the neutralisation of parasite variants associated with severe disease in semi-immune adults. Immunity to severe malaria can be acquired early in life (28) and so will require the rapid production of cross-reactive antibodies that recognise and eliminate group A and DC8 variants if these are sufficient to cause life-threatening disease. Whether antibodies can effectively neutralise these variants *in vivo* remains to be directly shown but the RNA-sequencing methodology that we developed for this study now allows transcriptional profiling of *P. falciparum* at ultra-low densities - indeed, we can spike just 2000 ring-stage parasites into 50ml whole blood (< 0.001% parasitaemia) and map their complete VSA profiles (Fig. 4). Applying this methodology to CHMI in an endemic setting would therefore allow switching & selection to be tracked under immune pressure in volunteers that can restrict and slow parasite replication.

One other intriguing finding that emerged from our analysis was that parasites from inflammatory volunteers express higher levels of *Pfck1* - a serine-threonine kinase with diverse roles in eukaryotic cells (73, 74). This enzyme has been shown to associate with the infected red cell membrane and is secreted into culture media suggesting the potential to regulate parasite-host interactions (75). In support of this idea, secreted CK1 enzymes have been shown to influence the virulence of *Leishmania major* (76) and *Toxoplasma gondii* (77), and may directly regulate type I interferon signaling (78). Our observation needs to be validated in a separate study but if confirmed would identify a possible mechanism through which malaria parasites may attempt to influence the host response to infection.

In summary, our study shows that *var* gene switching and selection does not occur during the first 6 cycles of blood-stage infection in naive hosts. No evidence was found to support the widely-held view that rapid expansion of parasites transcribing group A/DC8 *var* genes occurs *in vivo*, even in an inflammatory environment. Only one parasite gene *(Pfck1)* was differentially expressed in inflammatory volunteers. Hence, the diverse clinical outcomes of CHMI cannot easily be explained by variation in parasite gene expression and therefore mainly originate from human immune variation. The absence of switching/selection towards virulence-associated *var* genes was surprising, and suggests that a fundamental re-appraisal of one of the basic tenets of malaria pathogenesis should be considered.

## Methods

### Study participants and ethical approval

The fourteen volunteers recruited for blood challenge were infectivity controls in a phase I/IIa vaccine trial (VAC054) that took place at the Centre for Clinical Vaccinology and Tropical Medicine (CCVTM), Oxford (32). This study received ethical approval from the UK NHS Research Ethics Service (Oxfordshire Research Ethics Committee A, ref. 13/SC/0596) and the Western Institutional Review Board (WIRB) in the USA (ref. 20131985). The study was approved by the UK Medicines and Healthcare products Regulatory Agency (MHRA) (ref. 21584/0326/001–0001) and the trial was registered on ClinicalTrials.gov (NCT02044198). The five volunteers infected by mosquito bite were recruited at the same site and this study (VAC065) received ethical approval from the UK NHS Research Ethics Service (South Central Berkshire Research Ethics Committee, ref. 16/ SC/0261) and was approved by the UK MHRA (ref. 21584/0360/001–0001). The trial was registered on ClinicalTrials.gov (NCT02905019). Both trials were conducted according to the principles of the current revision of the Declaration of Helsinki 2008 and in full conformity with the ICH guidelines for Good Clinical Practice (GCP).

### Controlled human malaria infection

#### Blood challenge

details of inoculum preparation and assessment of parasite viability have been published previously (32). The fourteen volunteers (non-vaccinated infectivity controls) received an identical challenge by intravenous injection of 690 *P. falciparum* (clone 3D7) infected erythrocytes in 5ml of 0.9% saline. All inoculations were performed within 2 hours and 13 mins of inoculum preparation.

#### Mosquito challenge

CHMI was performed as previously described (79) using five infectious bites from *P. falciparum* (clone 3D7) infected *Anopheles stephensi* mosquitoes at the Alexander Fleming Building, Imperial College, London, UK. Infected mosquitoes were supplied by Jittawadee R. Murphy, Department of Entomology, Walter Reed Army Institute of Research, Washington, DC, USA.

### Monitoring volunteers

Blood samples were collected to measure parasitaemia by qPCR twice daily, starting 2-days post-infection for blood challenge and 6.5-days post-infection for mosquito challenge. In both trials, thick blood smears were also evaluated by experienced microscopists at each time-point. All reported clinical symptoms (pyrexia, fever, rigor, chills, sweats, headache, myalgia, arthralgia, back pain, fatigue, nausea, vomiting and diarrhoea) were recorded as adverse events and assigned a severity score: 1 - transient or mild discomfort (no medical intervention required); 2 - mild to moderate limitation in activity (no or minimal medical intervention required); 3 - marked or severe limitation in activity requiring assistance (may require medical intervention). A cumulative clinical score was then obtained by summing these adverse events across all time-points for each volunteer. At every clinic visit, staff at the CCVTM also assessed core temperature, heart rate and blood pressure. And on days –1, 6, 28 and 90 post-infection (plus diagnosis) full blood counts and blood biochemistry were performed at the Churchill and John Radcliffe Hospitals in Oxford, providing 5-part differential white cell counts and a quantitative assessment of electrolytes, urea, creatinine, bilirubin, alanine aminotransferase, alkaline phosphatase and albumin.

### Diagnostic criteria and drug treatment

Treatment with the artemether and lumefantrine combination drug Riamet® was prescribed to volunteers when two out of three diagnostic criteria were met: a positive thick blood smear and/or parasite density > 500 parasites ml^−1^ by qPCR and/or symptoms consistent with malaria.

### qPCR & parasite multiplication rate modeling

Blood was collected and prepared for qPCR analysis as previously described (32, 79). In brief, DNA was extracted from 0.5ml whole blood using the Qiagen Blood Mini Kit (as per manufacturer’s instructions) and 10% of each extraction was run in triplicate wells (equating to 150μl total blood volume). Previously published primers with TaqMan™ probes (5′ FAM-AAC AAT TGG AGG GCA AGNFQ-MGB 3′) (Applied Biosystems) were used to amplify the *P. falciparum* 18S ribosomal region and parasites ml^−1^ equivalent mean values were calculated using a defined plasmid standard curve and TaqMan™ absolute quantitation. Mean values below 20 parasites ml^−1^ (or values with only one positive replicate of the three tested) were classed as negative. Parasite multiplication rate (PMR) was then calculated using a linear model fitted to log10 transformed qPCR data, as previously described (79, 80). In the blood challenge model, fitted lines were constrained to pass through the known starting parasitemia, determined by the viability of the inoculum and a weight-based estimate of each volunteer’s total blood volume (32).

### Uninfected control volunteers

#### Blood challenge

four healthy adult volunteers were recruited at the University of Edinburgh to serve as uninfected controls for the blood challenge study. These volunteers had no history of malaria, and included 2 male and 2 female Caucasians with a mean age of 24 years (range 22–25). Written informed consent was given by all participants and the study was approved by the University of Edinburgh, School of Biological Sciences Ethical Review Committee (ref. arowe-0002 and pspence-0002). *Mosquito challenge*: one volunteer recruited to the VAC065 study was exposed to the bites of 5 infectious mosquitoes but did not develop measurable parasitaemia at any point during the 28-day study (limit of detection is 5 parasites ml^−1^). This volunteer therefore served as an internal uninfected control for the mosquito challenge study.

### Processing whole blood for host RNA and plasma

#### Blood challenge

for microarray analysis, 3ml venous blood was drawn directly into a Tempus™ Blood RNA Tube (Applied Biosystems), mixed and stored at –80°C. To obtain plasma, 14ml venous blood was drawn into lithium heparin vacutainers, transferred to Leucosep tubes (Greiner Bio-One) containing 15ml Lymphoprep (Axis Shield) and centrifuged at 1,000xg for 13 min. at room temperature. 2ml of the plasma fraction was collected, snap-frozen on dry ice and stored at –80°C. *Mosquito challenge*: for host RNA-sequencing, 6ml venous blood was drawn into EDTA-coated vacutainers, from which 1ml blood was removed and mixed with 2ml Tempus™ reagent; samples were stored at –80°C. To obtain platelet-depleted plasma, a further 1ml blood was removed from the vacutainer and given two spins; first at 1,000xg for 10 min. and then at 2,000xg for 15 min. (both at 4°C). After each spin, the plasma was transferred to a new tube (leaving behind any residual red cells or pelleted platelets) and finally the plasma was snap-frozen on dry ice and stored at –80°C.

### Host RNA extraction, quantification and quality control

RNA extraction was performed with the Tempus™ Spin RNA Isolation Kit (Applied Biosystems) according to the manufacturer’s instructions. The following modification was applied to mosquito challenge samples because of their reduced volume: after thawing, just 1ml PBS was added to each sample to maintain Tempus™ stabilizing reagent at the correct final concentration. Note that for all samples, a DNase treatment step using Absolute RNA wash solution (Applied Biosystems) was included for the removal of genomic DNA. Samples were quantified by nanodrop and RNA integrity was assessed on an Agilent 2100 Bioanalyzer using RNA 6000 nano chips. A RIN value above 7.0 was accepted as sufficiently high quality RNA for downstream steps.

### Host microarray and data processing (blood challenge)

Whole blood transcriptomics was undertaken with the Affymetrix GeneChip® Human Transcriptome Array 2.0 ST by Hologic Ltd. (Manchester, UK); samples were randomized to ensure unbiased sample-chip positioning. Raw array data in the form of compressed CEL files were processed in the R environment using Bioconductor packages (oligo, pd.hta.2.0, affy) to generate an HTAFeatureSet object (gene level). After visual inspection for QC purposes, raw and normalised (using the rma() function of the affy package or normalizeQuantiles() function from the limma Bioconductor package, as appropriate) versions of the intensities were retained for subsequent analyses. Array feature annotation was generated using the AnnotationForge package and the Bioconductor human reference database (org.Hs.eg.db).

#### Plotting the 100 protein-coding genes with highest variance

for each volunteer, a subset of the data was made encompassing all available time-points in chronological order. For each array feature, the variance of the intensities across the time-points was calculated using the *var*() function in R. The 100 protein-coding array features with the highest variance were then selected and for each of these features the median value of the intensities across all time-points was calculated and subtracted from the actual intensity values, resulting in a set of “deviation from median” values. These values were plotted using the heatmap.2() function in R and dendrograms, where shown, were calculated based on Euclidean distance measures of the input data (hclust() and dist() functions of the stats package).

#### Plotting the 517-gene superset

the lists of 100 protein-coding genes with highest variance in each of the 14 volunteers infected by blood challenge were merged to create a non-redundant set of 517 unique genes. Log2 transformed intensity values were then scaled with scale (scale = TRUE, center = TRUE), plotted with pheatmap and clustered using hclust and cutree in R, with k = 8.

#### PCA plots of the 517-gene superset

for each volunteer, a subset of the data was made encompassing all available time-points in chronological order. The log2 transformed intensity values of the 517 genes were normalised using the normalizeQuantiles()function from the limma Bioconductor package prior to principal component analysis using the prcomp() function of the stats Bioconductor package. Plots of the first two dimensions were generated using the standard plot() function of the graphics package. To measure the magnitude of each volunteer’s immune response we calculated the Euclidean distance travelled along principal component 1, as this accounted for almost all of the variance in every time-course. As every sample in a time-course was centred around zero we calculated the distance travelled for every volunteer (relative to their own average position through time) as the x-coordinate of the diagnosis sample.

#### Differential gene expression

analysis of differential gene expression was carried out using the limma package from Bioconductor, with eBayes correction for multiple testing; an adjusted p value below 0.05 was required for significance. This analysis was performed to quantify the number of differentially expressed genes between time-points for each group (unresponsive, inflammatory and suppressor). It was also carried out to quantify the number of differentially expressed genes between groups at a given time-point.

### Allocation of volunteers to groups after blood challenge

To be classified as responsive to blood challenge volunteers had to pass 2 sequential thresholds: first, for every volunteer the mean variance of their 50 most variable protein-coding genes was calculated. This figure had to be at least 1.5-fold higher than the mean of the four uninfected controls(i.e. 1.5 × 0.202 = **0.303**). Second, the distance travelled in each volunteer’s PCA plot of the 517-gene superset had to be greater than 2 standard deviations above the mean of the four uninfected controls(i.e. 2.67 + (2 × 1.102) = **4.874**). Volunteers had to pass both thresholds in order to be classified as responsive; those who failed one or both of these tests (v018, v012, v208 & v020) were classified as unresponsive. The remaining volunteers were then grouped into inflammatory (v016, v017, v013, v027, v026, v206, v106 & v024) or suppressor (v022 & v019) hosts based on their upregulation of clusters 2 & 4 or downregulation of clusters 1 & 3, respectively (see Fig. 1). Note that differential gene expression analysis (as detailed above) was used to confirm that the unresponsive group had zero differentially expressed genes at diagnosis (compared to baseline).

### Host RNA-sequencing and data processing (mosquito challenge)

Indexed sequencing libraries were prepared from 500ng RNA using the TruSeq Stranded mRNA Library kit (Illumina), according to the manufacturer’s instructions. Samples were pooled and sequenced on two separate lanes of the same Illumina HiSeq v4 flow cell to generate ∼ 33 million reads per sample (75bp paired-end reads). Note that globin depletion was not performed; instead sequencing was carried out to sufficient depth that the globin reads (accounting for no more than 20% of any given sample) could simply be discarded during data processing. Read counts per gene were quantified against the Gencode human v28 basic transcript sequences (81) using Kallisto v0.42.3 (82). The index was built using default parameters and quantification was performed with –t 12 –b 0; genes not expressed with at least 1 count per million in at least one sample were excluded. EdgeR (83) was used to identify highly dispersed genes (BCV > 0.4 and FDR < 0.1) for each volunteer’s time-course with the exact test. Next, the lists of highly dispersed genes in each of the 5 volunteers infected by mosquito bite were merged to create a non-redundant set of 226 unique genes, which were used for clustering with hclust in the pheatmap R package. This identified a single major cluster comprising 117 unique genes that was taken forward for downstream analyses (e.g. plotting the 117-gene superset and ClueGO). Note that to visualise and cluster the 226 unique genes and the 117-gene superset, read counts across the entire dataset were normalised using rlogs (blind = TRUE) in DESeq2 (84).

### Gene ontology analysis and networks (ClueGO)

#### Blood challenge

the list of 2,028 genes differentially expressed at diagnosis in the inflammatory group was imported into ClueGO v2.5.4 (33, 34). ClueGO identified the significantly enriched GO terms associated with these genes and placed them into a functionally organised non-redundant gene ontology network based on the following parameters: pvalue cutoff = 0.05; correction method used = Bonferroni step down; min. GO level = 5; max. GO level = 11; number of genes = 3; min. percentage = 5.0; GO fusion = true; sharing group percentage = 40.0; merge redundant groups with > 40.0% overlap; kappa score threshold = 0.4; and evidence codes used [All]. Each of the 34 functional groups was assigned a unique colour and a network was then generated using an edge-weighted spring-embedded layout based on kappa score. To provide an overview of the inflammatory response we then labelled the leading GO term in the top 12 functional groups, as follows: groups were first ordered by adj p value; starting with the most significant group we chose the level 4 GO term with the highest significance and made this the group name; we then moved on to the next most significant group and again chose the level 4 GO term with the highest significance as group name; we repeated this process until 12 groups had been named. Note that there were two exceptions to these rules: first, if there was not a level 4 GO term within a group then this group was excluded (to avoid naming groups that were too broad or specific in ascribing function); and second, if the most significant level 4 GO term was shared with a group already named then this GO term was excluded and the group was instead named after the next most significant level 4 GO term (to avoid redundancy).

#### Mosquito challenge

the 117-gene superset was imported into ClueGO to identify the significant GO terms associated with these genes. This was carried out exactly as described for blood challenge with the exception of the following parameter changes: number of genes = 2; min. percentage = 4.0; merge redundant groups with > 50.0% overlap.

### Quantification of cytokines & chemokines in plasma

Interferon alpha (IFNα), Interferon gamma (IFNγ), CXCL9 (MIG) and CXCL10 (IP-10) were quantified in plasma as part of a custom-design multiplex assay from BioLegend (LegendPlex™). Samples were run in duplicate and the assay was performed according to the manufacturer’s instructions using low protein-binding filter-bottom plates (Merck). Beads were acquired on a BD LSRFortessa™ running FACSDiva™ software and data were analysed with LEGENDPlex™ analysis software. For blood challenge, data are presented as fold-change relative to pre-infection samples; when analytes were undetectable samples were simply assigned the largest integer below the limit of detection to allow normalisation and graphing of all time-points.

### Quantification of Angiopoietin-2 in plasma

Angiopoietin-2 was measured in plasma at pre-infection and diagnosis time-points using the Human Angiopoietin-2 Quantikine ELISA kit from R&D Systems. Samples were run in duplicate and the assay performed as per the manufacturer’s instructions. Optical density (OD) was determined on a Multiskan Ascent plate reader by subtracting measurements taken at 570nm wavelength from readings at 450nm. All samples and standards were background-corrected by subtracting the mean OD measured in blank wells containing only assay diluent. A 7-point standard curve was then constructed by plotting the mean absorbance of each standard against known concentration and using regression analysis to draw a best fit line (R^2^ = 0.998). Data are presented as mean concentration of the duplicate samples.

### Detection of anti-Cytomegalovirus IgG in plasma

Cytomegalovirus (CMV) seropositivity was assessed in all volunteers before CHMI using an anti-CMV IgG Human ELISA Kit (Abcam). Samples were run in duplicate and OD was determined on a Multiskan Ascent plate reader by subtracting measurements taken at 620nm wavelength from readings at 450nm. All samples and controls were background-corrected by subtracting the mean OD measured in substrate blank wells. Samples were scored positive if their absorbance value was > 10% above the mean absorbance value of the CMV IgG cut-off control. This was calculated by converting OD values to Standard Units using the following equation: mean sample absorbance value × 10 / mean absorbance value of the cut-off control. Samples with < 9 standard units were classified as CMV-negative whereas samples with > 11 standard units were classified as CMV-positive.

### LC-MS-based analysis of plasma metabolites

Samples were prepared by diluting 25μl of plasma in 1,000μl of 4°C (HLPC-grade) chloroform/ methanol/water at a 1:2:1 ratio. Samples were then vortexed for 5 min. and spun at 13,000xg for 3 min. at 4°C. After centrifugation, the supernatant was collected and stored in cryovials at –80°C ready for downstream processing. LC-MS-based metabolomics analyses were conducted at Glasgow Polyomics. Hydrophilic interaction liquid chromatography (HILIC) was performed on a Dionex UltiMate 3000 RSLC system (Thermo Fisher Scientific, UK) using a ZIC-pHILIC column (150mm × 4.6mm, 5μm column from Merck Sequant). For each sample, a 10μl volume was injected into the system. Samples were maintained at 5°C prior to injection. The column was kept at 30°C throughout the analysis and samples were eluted with a linear gradient (from 20% 20mM ammonium carbonate in water (A) and 80% acetonitrile (B) to 80% A and 20% B) at a flowrate of 300 μl/min over 24 min. Mass spectrometry analysis was performed on a Thermo Orbitrap QExactive operated in polarity-switching ionization mode with the following parameters: resolution of 70,000; automated gain control of 106; m/z range of 70–1,050; sheath gas flowrate of 40 arbitrary units (au); auxiliary gas flowrate of 5 au; sweep gas flowrate of 1 au; probe temperature of 150°C and capillary temperature of 320°C. The parameters for positive mode ionisation were as follows: source voltage +3.8 kV, S-Lens RF Level 30.00, S-Lens Voltage –25.00 V, Skimmer Voltage –15.00 V, Inject Flatapole Offset –8.00 V, Bent Flatapole DC –6.00 V. For negative mode ionisation, a source voltage of –3.8 kV was used. Volunteer samples were run alongside three mixes of authentic standards (total number: 248) involved in various metabolic pathways to facilitate metabolite identification based on exact mass and retention time. Fragmentation of the top 20 ions was performed with the following parameters: collision energy: 25%; isolation window: 1.2; dynamic exclusion after 1 time; exclusion duration: 6 seconds; exclude isotopes: true; and minimum intensity: 5000. LC-MS raw data were processed with IDEOM (using default parameters (85)), which uses the XCMS (86) and mzMatch (87) software in the R environment. The levels of reliability of the spectral assignment to metabolites, as defined by the Metabolomics Standard Initiative (88), are as follows: ‘MSI:1 (identified metabolites)’ - high resolution mass (3 ppm) and retention time (5%) matched to an authentic standard; and ‘MSI:2 (putatively annotated compounds)’ - high resolution mass matched to a public library (3 ppm). Xcalibur™ software (Thermo Fisher Scientific) was used to generate fragmentation spectra, which were further exported into mzCloud™ to search for compounds in the database with matching spectra.

### Metabolomics data processing & analysis

Metabolite intensities were log2 transformed and intensity ratios were calculated as the fold-change in metabolite abundance at day 8 or diagnosis relative to pre-infection. A pairwise comparison (t-test) between post- and pre-infection samples was then carried out for every metabolite and p values were adjusted for multiple testing by applying a false discovery rate (FDR). Differentially abundant metabolites were called using an FDR threshold of 0.1 and an intensity ratio equivalent to a 1.5-fold change in abundance. A volcano plot showing the intensity ratios and FDR-corrected p values of all metabolites was generated using ggplot2. Next, the differentially abundant metabolites were range-scaled according to van den Berg *et al* (45) and a heatmap of range-scaled intensity values was generated using the gplots R package. Here range-scaling is used to make every metabolite equally important by scaling the intensity of each metabolite relative to its own biological range across all samples.

### Isolation of parasites for *ex vivo* RNA-sequencing

#### Blood challenge

50ml whole blood was drawn into lithium heparin vacutainers at diagnosis (immediately before drug treatment) and white cells were removed by passing the blood through a leucoflex LXT filter (Macopharma). The flow-through was collected into a 250ml centrifuge bottle (with silicone O-ring) and the filter was washed with 200ml PBS to maximise recovery of infected red cells. The flow-through was spun at 1,000xg for 10 min. at room temperature with brake off and the supernatant carefully aspirated (being careful not to disturb the pelleted red cells). The red cells were gently agitated to resuspend and then lysed by addition of 200ml ice-cold saponin at 0.015% in PBS (w/v). The cell suspension was incubated on ice for 10min. and then spun at 15,000xg for 10 min. at 4°C with brake off to pellet the free parasites. The supernatant was slowly aspirated being very careful not to disturb the pelleted parasites, which were then gently resuspended in 1ml ice-cold PBS and transferred to a 1.5ml tube. The parasites were pelleted by centrifugation at 16,000xg for 5 min. at 4°C in a microfuge and two sequential washes were performed with 1ml ice-cold PBS to remove free globin. After the final spin, the parasite pellet was resuspended in 1ml TRIzol, incubated in a 37°C water bath for 5 min. and snap-frozen on dry ice. All samples were stored at –80°C prior to RNA extraction.

#### Mosquito challenge

parasites were isolated from 20ml whole blood drawn on days 9, 10 and 11 post-infection according to the blood challenge protocol, with the following modifications. Red cells were lysed by addition of 80ml ice-cold saponin at 0.0075% in PBS (w/v); after the 10 min. incubation on ice, free parasites were then pelleted by centrifugation at 18,000xg for 20min. at 4°C with brake off.

#### Inoculum

two samples of the inoculum used for blood challenge were prepared to create biological replicates for RNA-sequencing. For each sample, 1ml of the pre-diluted inoculum (estimated to contain 1000 parasites ml^−1^ in 0.9% saline) was transferred to a 1.5ml tube; infected red cells were pelleted by centrifugation at 1000xg for 5 min. at room temperature and (after removal of the supernatant) resuspended in 1ml TRIzol. The samples were carefully mixed, incubated in a 37°C water bath for 5 min. and then snap-frozen on dry ice.

#### In vitro cultured ring-stage parasites

two mock samples were prepared using *in vitro* cultured parasites (clone 3D7) to test the sensitivity of our ultra-low input methodology for parasite RNA-sequencing. Briefly, parasites were cultured and synchronised to ring-stage using 5% sorbitol, and the first mock sample was prepared by spiking 2000 parasites into 50ml whole blood (40 parasites ml^−1^). The second mock sample was prepared by spiking 8000 parasites into 20ml whole blood (400 parasites ml^−1^). These parasite densities were chosen to represent the likely burden in the first and second blood cycle after liver egress in volunteers infected by mosquito bite (7). Parasites were then isolated from these mock samples according to the mosquito challenge protocol outlined above.

### Parasite RNA extraction and depletion of globin & ribosomal RNA

Samples were thawed at room temperature and 200μl bromochloropropane was added; samples were then vortexed, incubated for 3 min. at room temperature and centrifuged at 12,000xg for 15 min. at 4°C. 500μl of the aqueous phase was transferred to a 1.5ml tube and diluted 1:1 with absolute ethanol. RNA purification was then carried out using the RNA Clean and Concentrator™ kit, as per the manufacturer’s instructions (Zymo Research). RNA was eluted in 14μl DEPC-treated water and quantified by nanodrop; quality was assessed using the Agilent 2100 Bioanalyzer with RNA 6000 pico chips. A RIN value above 7.0 was accepted as sufficiently high quality RNA for downstream steps. Samples were then DNase-treated and depleted of globin & ribosomal RNA with the Globin-Zero® Gold kit (Illumina), according to the manufacturer’s instructions. For samples with less than 1μg RNA (the recommended starting material) reagent volumes were downscaled for low-input methodology (as described in the kit). After removal of globin and ribosomal RNA, the remaining RNA was precipitated overnight at 4°C in 600μl absolute ethanol, 18μl sodium acetate (3M) and 2μl linear acrylamide (5mg/ml). The RNA was then pelleted by centrifugation at 12,000xg for 20 min. at 4°C and washed two times with 75% ethanol. Samples were air-dried for 5 min. and the RNA was dissolved in 12μl DEPC-treated water by heating to 65°C for 5 min. RNA was stored at –80°C prior to cDNA synthesis and amplification.

### Parasite RNA-sequencing and data processing

#### Blood challenge

double-stranded cDNA was synthesized from 2μl parasite RNA using the Smart-Seq2 protocol described by Picelli *et al* (89) and modified for use with low-input *P. falciparum* RNA (90). cDNA was amplified with 25 cycles of PCR using the KAPA HiFi reaction mix and ISPCR primer, and purified with Agencourt AMPure XP beads at a 1:1 DNA to bead ratio. Indexed sequencing libraries were constructed from 500ng of cDNA with no further PCR amplification. cDNA was sheared to 400–600bp with a Covaris sonicator and libraries prepared using the NEB NEXTflex library kit, according to the manufacturer’s instructions. Libraries were pooled and run on two separate lanes of distinct Illumina HiSeq v4 flowcells to produce 75bp paired-end reads. Read counts per gene were quantified against the *P. falciparum* v3 transcript sequences (91) using Kallisto v0.42.3 (82). The index was built using default parameters and quantification was performed with –t 12 –b 0. *Mosquito challenge*: double-stranded cDNA was synthesized and amplified exactly as described for blood challenge, with the exception that the number of PCR cycles was reduced to 20. Indexed sequencing libraries were prepared from 1ng cDNA with the Nextera XT library kit (Illumina), as per the manufacturer’s instructions. 1μl of each undiluted library was run on the Agilent bioanalyzer high sensitivity DNA chip for quantification and quality assessment, and all libaries were pooled and run on a single lane of an Illumina Hiseq v4 flowcell. Read counts per gene were quantified against the *P. falciparum* v3 transcript sequences using Kallisto v0.42.3, as above. The index was built using default parameters and quantification was performed with –t 12 –b 0.

### Analysis of parasite *var* and *rifin* expression

For each volunteer, the read counts were combined from days 9, 10 and 11 after mosquito challenge. Data from the mosquito and blood challenge studies were then merged and rlog values (blind = TRUE) were calculated across the entire dataset using DESeq2 (84); these values were used to generate heatmaps of expression intensity. Sub-family classifications for the 62 *var* genes were determined from PlasmoDB ((92) based on (93)) and 158 *rifin* genes were identified from the same database. Heatmaps were drawn using pheatmap, clustered with hclust. To generate pie charts of *var* gene sub-family expression we summed FPKM-normalised (Fragments Per Kilobase per Million mapped reads) read counts for each *var* gene sub-family for each volunteer. Genes with a summed FPKM of < 1 in any given sample were excluded for that sample and plots were drawn using matplotlib in Python. To examine reads mapping across *var* gene introns, we followed the protocol outlined in Reid *et al* (90). Briefly, we generated a HISAT2 (v2.0.0-beta) index of the *P. falciparum* v3 genome sequence using default parameters and mapped reads with parameters –-max-intronlen 5000 –p 12 (94). We identified reads overlapping annotated *var* genes using bedtools intersect (95). We then selected reads with an N in the CIGAR string, indicating a split read. We counted only those reads that were split exactly over the intron of a *var* gene. We called expression for a *var* gene where there were at least two reads mapping over the intron; read counts were then converted to logs of Counts Per Million (CPM) using the total number of reads mapping to all other features and the heatmap was drawn using pheatmap.

### Calling differentially expressed parasite genes

To determine whether any parasite genes were differentially expressed between volunteers with different clinical outcomes after blood challenge, we used DESeq2 (84) to compare parasite gene expression profiles from inflammatory volunteers (n = 8) versus the rest (unresponsive and suppressor, n = 4). In the same way, we used DESeq2 to compare parasite gene expression profiles between the inoculum samples (n = 2) and all volunteer samples (n = 12) after blood challenge.

### Accession numbers

Blood challenge human microarray dataset (NCBI GEO): GSE132050

Mosquito challenge human RNA-sequencing dataset (EBI ArrayExpress): EGAS00001003766 and EGAD00001005790

Blood challenge metabolomics dataset (MetaboLights): MTBLS1188

Parasite RNA-sequencing dataset (blood and mosquito challenge) (EBI ENA): ERP116360

## Data Availability

All datasets are publicly available using the accession numbers below:
Blood challenge human microarray dataset (NCBI GEO): GSE132050
Mosquito challenge human RNA-sequencing dataset (EBI ArrayExpress): EGAS00001003766 and EGAD00001005790
Blood challenge metabolomics dataset (MetaboLights): MTBLS1188
Parasite RNA-sequencing dataset (blood and mosquito challenge) (EBI ENA): ERP116360

## Acknowledgements

We are grateful to Julie Furze, Sean Elias, Katie Ewer, the VAC054 study team and the VAC065 study team (Jenner Institute Laboratories and the Centre for Clinical Vaccinology and Tropical Medicine, University of Oxford) for assistance and also for access to samples from these CHMI clinical trials (originally funded by the PATH Malaria Vaccine Initiative, US Agency for International Development (USAID) and the European Union Seventh Framework Programme (FP7/2007–2013) under the grant agreement for MultiMalVax (number 305282)). We also thank all of the clinical trial volunteers and blood donors who participated in this study.

